# The post-viral GPNMB^+^ immune niche persists in long-term Covid, asthma, and COPD

**DOI:** 10.1101/2024.08.27.24312640

**Authors:** Kangyun Wu, Yong Zhang, Huiqing Yin-DeClue, Kelly Sun, Dailing Mao, Erika C. Crouch, Derek E. Byers, Michael J. Holtzman

## Abstract

Epithelial injury calls for a regenerative response from a coordinated network of epithelial stem cells and immune cells. Defining this network is key to preserving the repair process for acute resolution, but also for preventing a remodeling process with chronic dysfunction. We recently identified an immune niche for basal-epithelial stem cells using mouse models of injury after respiratory viral infection. Niche function depended on an early sentinel population of monocyte-derived dendritic cells (moDCs) that provided ligand GPNMB to basal-ESC receptor CD44 for reprogramming towards chronic lung disease. These same cell and molecular control points worked directly in mouse and human basal-ESC organoids, but the findings were not yet validated in vivo in human disease. Further, persistence of GPNMB expression in moDCs and M2-macrophages in mouse models suggested utility as a long-term disease biomarker in humans. Here we show increased expression of GPNMB localized to moDC-macrophage populations in lung tissue samples from long-term Covid, asthma, and COPD. The findings thereby provide initial evidence of a persistent and correctable pathway from acute injury to chronic disease with implications for cellular reprogramming and inflammatory memory.

**New and noteworthy:** Recent work indicates that a sentinel immune niche provides GPNMB to epithelial stem cells to drive structural remodeling and disease as exemplified by the response to respiratory viral injury. The present study provides initial evidence that this niche can be detected in humans in the context of comparable diseases (long-term Covid, asthma, and COPD) also linked to viral infection. The results support a persistent mechanism for inflammatory disease that might be correctable with GPNMB blockade directly or indirectly through related signaling pathways.

## Introduction

Epithelial barriers are carefully programmed for primordial defense and repair in response to injuries from infectious and other toxic agents. In that context, one of the most common forms of epithelial injury derives from respiratory viral infection, including childhood outbreaks of RSV and HEV-D68 and pandemics of influenza virus and coronavirus (1, 2). In each case, the host goal is to wall off infection and restore integrity at the barrier site. A key step in this repair process is the growth and differentiation of epithelial stem cells and the coordinated actions of immune cells designed to clear infectious agents. However, based on viral and host factors, the normal program for recovery can be skewed to an ongoing response that results in structural remodeling and long-term post-viral lung disease (PVLD) that can manifest as long-Covid, post-influenza sequelae and related virus-triggered diseases such as asthma and COPD (3-6). In fact, basal-epithelial stem cells (basal-ESCs) represent a stereotyped feature of the epithelial barrier program. In experimental models, this cell population can be reprogrammed for hyperplasia and metaplasia that disrupts lung function after native Sendai virus (SeV) or adapted influenza A virus (IAV) infections (7-9). Similar activation of basal cell growth is found in Covid-19 (6, 10) that might be linked to asthma exacerbation (11). Defining and correcting a renewable stem cell component with the capacity for inflammatory memory is entry point for a disease-modifying therapy. However, the molecular basis for excess growth and immune activation still needed to be defined as a basis for precisely targeted correction.

To address these issues, we recently identified a sentinel immune niche for basal-ESC reprogramming in mouse models of epithelial injury after respiratory viral infection. Niche function depended on monocyte-derived dendritic cell (moDC) recruitment and then production of ligand glycoprotein nometastatic melanoma B (GPNMB) for delivery to receptor CD44 on basal-ESCs. This ligand-receptor interaction could be antibody-blocked early after infection (5-12 d) to prevent the subsequent reprogramming and PVLD that developed later after infection (49 d). These same moDC and GPNMB-CD44 control points worked directly in mouse and human basal-ESC organoids, but the findings were not yet extended to studies of human disease conditions. Further, persistence of GPNMB expression in moDCs and then M2-macrophages after clearance of infection suggested utility as a long-term biomarker for chronic lung disease. In line with these concepts, we show here that expression of GPNMB can also be localized to lung moDCs and macrophages in situ in humans using post-mortem tissues from long-term Covid, asthma, and COPD. The results thereby provide initial evidence of methods to stratify and modify post-injury disease in the lung and perhaps other sites of epithelial injury.

## Results and Discussion

To determine whether findings in experimental animal and human models translate to similar characteristics in human lung disease, we engaged a tissue registry of human lung samples constructed and validated as described previously (3, 6, 12, 13). For long-term Covid samples, human lung tissue was obtained from a series of consecutive autopsies performed from April-August 2020 at 27-51 d after onset of infectious illness (6). For asthma, COPD, and non-disease control samples, lung tissue was obtained from a Tissue Registry for Advanced Lung Disease that contains whole lung explants harvested but not used for lung transplantation and from a tissue procurement service (IIAM, Edison, NJ) (3, 12, 13). For the present experiments, the clinical characteristics of lung tissue donors is summarized in **Table 1**, recognizing that full characteristics were unavailable for some donors that provided tissues collected post-mortem.

**Table 1:**
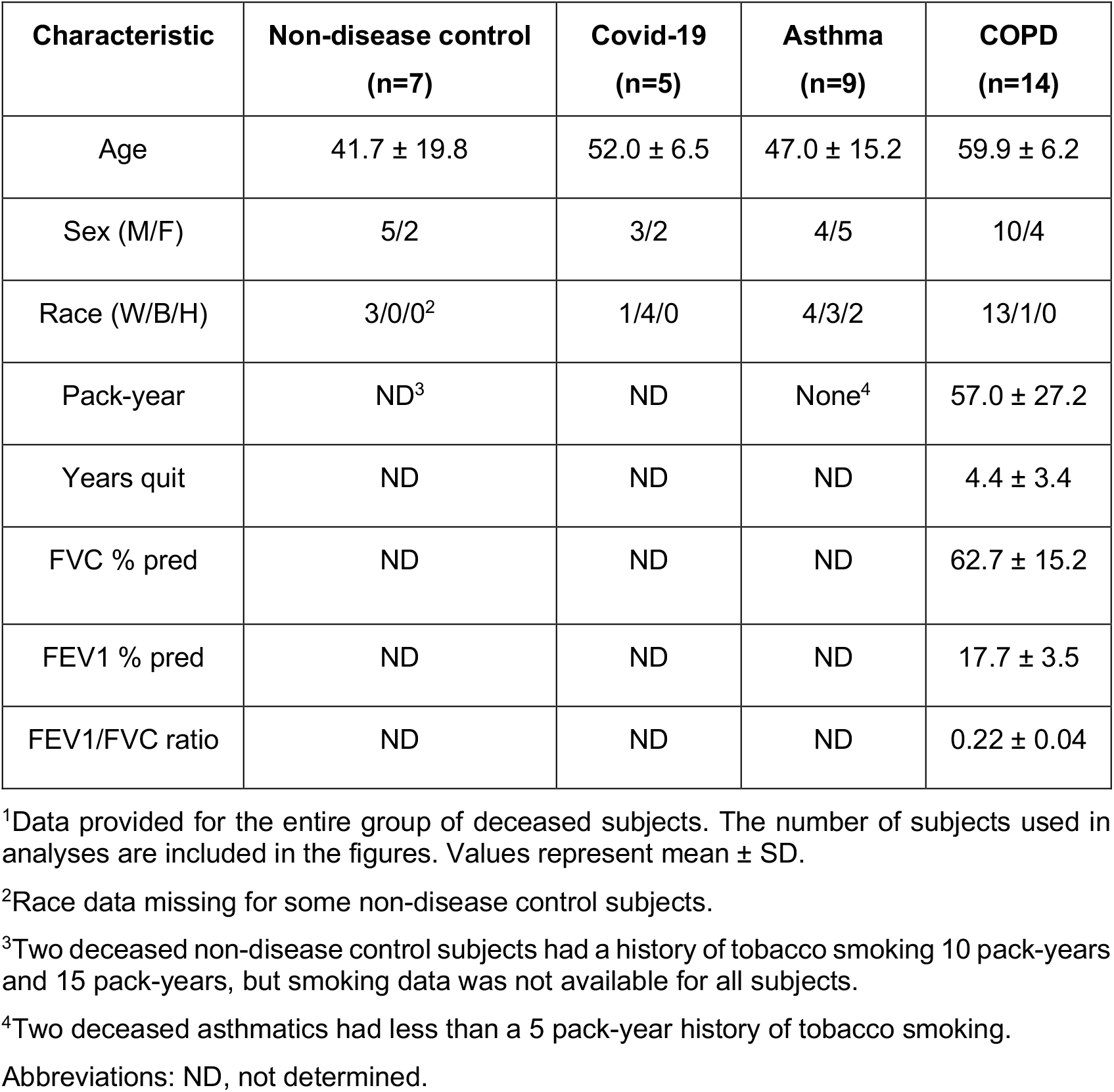
Clinical characteristics of tissue sample groups.

The primary endpoints for study were the site and level of expression for GPNMB in disease versus non-disease control conditions. Detection methods were the same those applied to studies of GPNMB expression in mouse models of PVLD (14) to allow for comparison across experimental and clinical conditions. Using this approach, immunostaining of lung tissue sections showed expression of GPNMB localized to CD11c^+^ and CD68^+^ cells with moDC and macrophage morphology in long-term Covid, asthma, and COPD (**Figure 1A**). Quantitative morphology demonstrated that levels of GPNMB expression were significantly increased in each disease condition compared to non-disease control (**Figure 1B**). In concert with GPNMB expression, immunostaining for CD44 was also localized (although not exclusively) to basal epithelial cells under disease and control conditions (**Figure 1A**).

**Figure 1.**
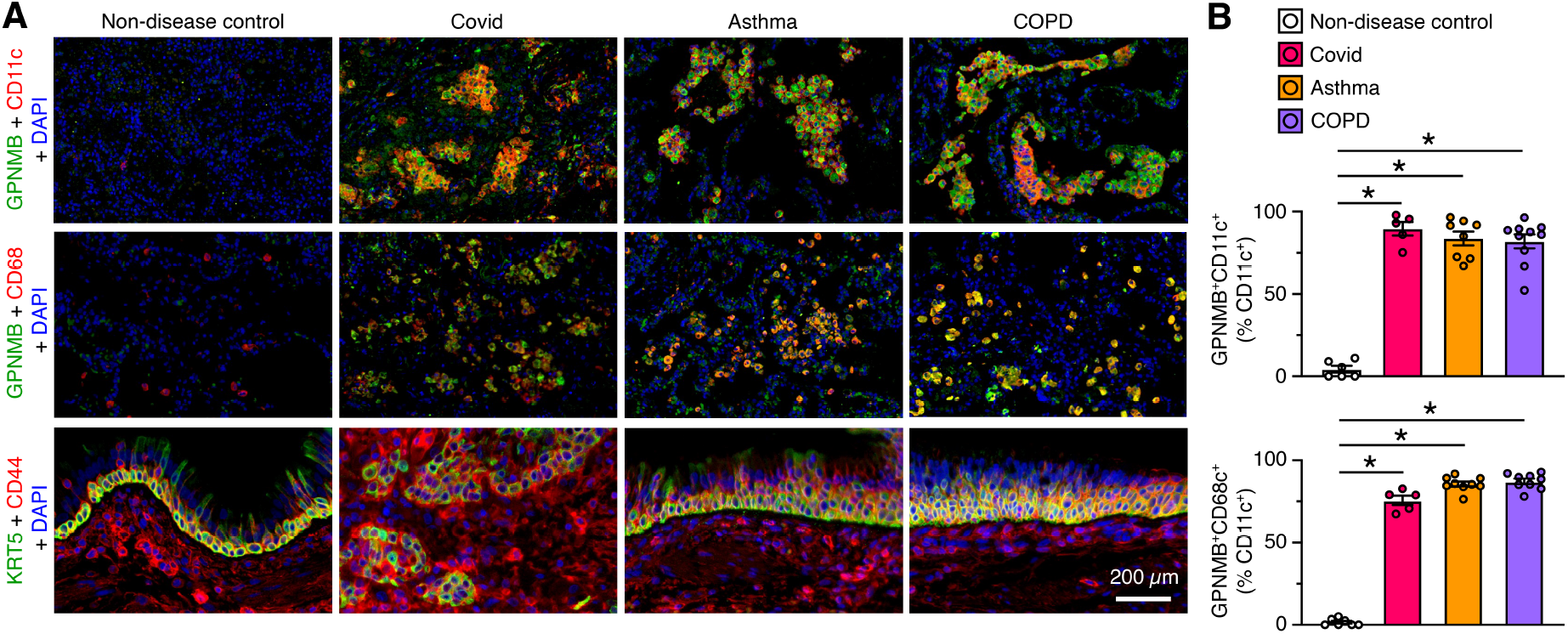
GPNMB and CD44 expression in chronic lung disease. **A**, Representative immunostaining for GPNMB plus CD11c or CD68 and KRT5 plus CD44 with DAPI counterstaining in lung sections from non-disease control (n=5-6), Covid (n=5), asthma (n=8-9), and COPD (n=9-10) subjects. **B**, Quantitation of immunostaining from (A). Values represent mean ± SEM. \**P* <0.05 by ANOVA and Tukey correction for multiple comparisons. No significant differences were found among disease conditions.

Together, the present findings provide initial validation for comparable cell and molecular components in experimental models and clinical samples of chronic lung disease. In the experimental setting, GPNMB expression is significantly increased and localized to moDCs and macrophages in concert with basal-ESC hyperplasia/metaplasia and immune activation that feed-forward to promote additional immune cell infiltration. The relatively prolonged time course predicted a comparable GPNMB^+^ moDC and macrophage signature in chronic lung disease even long after any previous injury. Indeed, that appears to be the case given the prominence of GPNMB^+^CD68^+^ macrophages found in clinical samples of lung tissue in long-term Covid-19, asthma, and COPD. Thus, the present findings are comparable to the later phase (21-49 d after infection) of the viral mouse model wherein similar GPNMB expression can be localized to M2 macrophages (14).

The present data suggests the presence of a persistent GPNMB signal in disease, raising the question of mechanism for how this signal remains active. Certainly having the signal in long-lived cells (in this case moDCs and macrophages but in other cases basal-ESCs) is a contributing factor, but this might not be sufficient to explain long-term persistence over years. In that regard, beneficial instructions for host defense, repair, and inflammatory memory after viral infection (15) or other injuries might instead manifest as long-term, epigenetic reprogramming for inflammatory disease. Passing these instructions in renewable cell populations would provide even longer reprogramming towards disease. In this case, GPNMB and related biomarkers could provide guidance for detecting and correcting this type of disease. Thus, correlation with GPNMB signaling partners as recently identified (14, 16, 17) will also be instructive. Extending the present studies of post-mortem tissues to the clinical precision of a planned patient enrollment and comprehensive survey study will also better define the differences between non-disease and disease conditions. The present and pending information should provide significant practical value given the potential for correcting GPNMB and related signaling activities as a mechanism to modify post-injury disease.

## Materials and Methods

### Human clinical samples

For Covid-19 samples, human lung tissue was obtained from a series of consecutive autopsies performed from April-August 2020 at Barnes-Jewish Hospital as described previously (6). For asthma, COPD, and non-disease control samples, lung tissue was obtained from a Tissue Registry for Advanced Lung Disease that contains whole lung explants harvested but not used for lung transplantation and from a tissue procurement service (IIAM, Edison, NJ) as described previously (3, 6, 12, 13). Human studies were conducted with protocols approved by the Washington University (St. Louis, MO) Institutional Review Board and U.S. Army Medical Research and Development Command (USAMRDC) Office of Research Protections.

### Histology and immunostaining

Lung tissue was fixed with 10% formalin, embedded in paraffin, cut into 5-μm sections and adhered to charged slides. Sections were stained with PAS and hematoxylin as described previously (9, 18). For immunostaining, sections were deparaffinized in Fisherbrand® CitriSolv® (Fisher), hydrated, and heat-treated with antigen unmasking solution (Vector Laboratories, Inc). Immunostaining was performed with the commercially available primary antibodies as listed in **Table 2**. Primary Abs were detected with secondary Abs labeled with Alexa Fluor 488 (Thermo Fisher Scientific) or Alexa Fluor 594 (Thermo Fisher Scientific) followed by DAPI counterstaining. Slides were imaged by light microscopy using a Leica DM5000 B and by immunofluorescent microscopy using an Olympus BX51, and staining was quantified in whole lung sections using a NanoZoomer S60 slide scanner (Hamamatsu) and ImageJ software as described previously (9, 18).

**Table 2.**
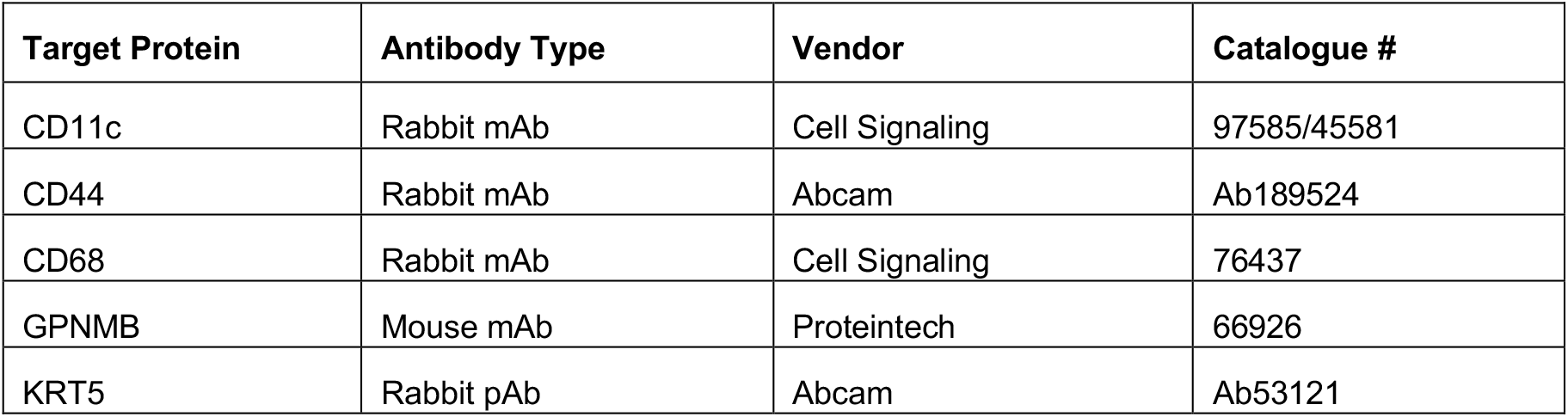
Antibodies for human tissue immunostaining.

| **Target Protein** | **Antibody Type** | **Vendor** | **Catalogue #** |
| --- | --- | --- | --- |
| CD11c | Rabbit mAb | Cell Signaling | 97585/45581 |
| CD44 | Rabbit mAb | Abcam | Ab189524 |
| CD68 | Rabbit mAb | Cell Signaling | 76437 |
| GPNMB | Mouse mAb | Proteintech | 66926 |
| KRT5 | Rabbit pAb | Abcam | Ab53121 |

### Statistical analysis

All data presented in bar-graph formats were expressed as mean ± SEM or SD as indicated. For this data, statistical differences between means for sample conditions were assessed using one-way analysis of variance (ANOVA) with Tukey correction for multiple comparisons. For all data, significance threshold was set at *P*<0.05. The number of human subjects for each condition is defined in the legend for **Figure 1**. These subjects were selected at random from the total group of subjects shown in **Table 1**.

## Data Availability

All data produced in the present work are contained in the manuscript.

## Abbreviations used in this article

basal-ESC: basal-epithelial stem cell
COPD: chronic obstructive pulmonary disease
Covid-19: coronavirus disease of 2019
GPNMB: glycoprotein nometastatic melanoma B
(moDC): monocytederived dendritic cell
PVLD: post-viral lung disease.

## Acknowledgments

We thank the Pulmonary Morphology Core and Anatomic and Molecular Pathology Core Labs for outstanding technical support.

## Funding

This study was supported by grants from the National Institutes of Health (National Heart, Lung, and Blood Institute UH2-HL123429, R35-HL145242, and STTR R41-HL149523, National Institute of Allergy and Infectious Diseases R01-AI130591, Department of Defense TTDA W81XWH2010603 and W81XWH2210281, and Harrington Discovery Institute.

## Disclosures

MJH is the Founder of NuPeak Therapeutics, Inc. KW, YZ, AGR, and MJH are inventors on a patent for MAPK inhibitors and methods of use thereof. MJH, KW, and YZ are inventors on a provisional patent for Methods of use for GPNMB-CD44 blockade in chronic respiratory disease.

## Author contributions

K.W. organized and performed experiments, Y.Z. organized experiments, H.Y-D. performed immunostaining; D.M. performed experiments, K.S. performed experiments; E.C.C. identified and analyzed autopsy samples; D.E.B. obtained and libraried donor samples; and M. J. H. directed the project and wrote the manuscript.

## Notes

### Author Declarations

Human studies were conducted with protocols approved by the Washington University (St. Louis, MO) Institutional Review Board and U.S. Army Medical Research and Development Command (USAMRDC) Office of Research Protections.

## References

1. Bautista E, Chotpitayasunondh T, Gao Z, Harper SA, Shaw M, Uyeki TM, Zaki SR, Hayden FG, Hui DS, Kettner JD, Kumar A, Lim M, Shindo N, Penn C, and Nicholson KG. Clinical aspects of pandemic 2009 influenza A (H1N1) virus infection. N Engl J Med. 2010;362:1708–19.

2. Yang X, Yu Y, Xu J, Shu HB, Xia J, Liu H, Wu Y, Zhang L, Yu Z, Fang M, Yu T, Wang Y, Pan S, Zou X, Yuan S, and Shang Y. Clinical course and outcomes of clinically ill patients with SARS-CoV-2 pneumonia in Wuhan, China: a single-centered, retrospective, observational study. Lancet Respir Med. 2020;8:475–81.

3. Alevy Y, Patel AC, Romero AG, Patel DA, Tucker J, Roswit WT, Miller CA, Heier RF, Byers DE, Brett TJ, and Holtzman MJ. IL-13–induced airway mucus production is attenuated by MAPK13 inhibition. J Clin Invest. 2012;122:4555–68.

4. Kim EY, Battaile JT, Patel AC, You Y, Agapov E, Grayson MH, Benoit LA, Byers DE, Alevy Y, Tucker J, Swanson S, Tidwell R, Tyner JW, Morton JD, Castro M, Polineni D, Patterson GA, Schwendener RA, Allard JD, Peltz G, and Holtzman MJ. Persistent activation of an innate immune response translates respiratory viral infection into chronic lung disease. Nat Med. 2008;14:633–40.

5. Byers DE, Alexander-Brett J, Patel AC, Agapov E, Dang-Vu G, Jin X, Wu K, You Y, Alevy YG, Girard J-P, Stappenbeck TS, Patterson GA, Pierce RA, Brody SL, and Holtzman MJ. Long-term IL-33-producing epithelial progenitor cells in chronic obstructive lung disease. J Clin Invest. 2013;123:3967–82.

6. Wu K, Zhang Y, Yin Declue H, Austin SR, Byers DE, Crouch EC, and Holtzman MJ. Lung remodeling regions in long-term coronavirus disease 2019 feature basal epithelial cell reprogramming. Am J Pathol 2023;193:680–9.

7. Zuo W, Zhang T, Zheng D, Wu A, Guan SP, Liew A-A, Yamamoto Y, Wang X, Lim SJ, Vincent M, Lessard M, P. Cc, Xian W, and McKeon F. p63+Krt5+ distal airway stem cells are essential for lung regeneration. Nature. 2015;517:616–20.

8. Vaughan AE, Brumwell AN, Xi Y, Gotts JE, Brownfield DG, Treutlein B, Tan K, Tan V, Liu FC, Looney MR, Matthay MA, Rock JR, and Chapman HA. Lineage-negative progenitors mobilize to regenerate lung epithelium after major injury. Nature. 2015;517:621–5.

9. Wu K, Kamimoto K, Zhang Y, Yang K, Keeler SP, Gerovac BJ, Agapov EV, Austin SP, Yantis J, Gissy KA, Byers DE, Alexander-Brett J, Hoffmann CM, Wallace M, Hughes ME, Morris SA, and Holtzman MJ. Basal-epithelial stem cells cross an alarmin checkpoint for post-viral lung disease. J Clin Invest. 2021;131:e149336.

10. Delorey TM, Ziegler CGK, Heimberg G, Normand R, Yang Y, Segerstolpe A, Abbonddanza D, Fleming SJ, Subramanian A, Montoro DT, Jagadeesh KA, Dey KD, Sen P, Slyper M, Pita-Juarez YH, and Phillips D. COVID-19 tissue atlases reveal SARS-CoV-2 pathology and cellular targets. Nature. 2021;595:107–13.

11. Lee H, Kim B-G, Jeong CY, Park DW, Park TS, Moon J-Y, Kim T-H, Sohn JW, Yoon HJ, Kim JS, and Kim S-H. Long-term impacts of COVID-19 on severe exacerbation and mortality in adult asthma: a nationwide populations-based cohort study. J Allergy Clin Immunol Pract. 2024;12:1783–93.

12. Deslee G, Woods J, Moore C, Conradi S, Gierada D, Atkinson J, Battaile J, Liu L, Patterson A, Adair-Kirk T, Holtzman M, and Pierce R. Oxidative damage to nucleic acids in severe emphysema. Chest. 2009;135:965–74.

13. Byers DE, Wu K, Dang-Vu G, Jin X, Agapov E, Zhang X, Battaile JT, Schechtman KB, Yusen R, Pierce RA, and Holtzman MJ. Triggering receptor expressed on myeloid cells-2 (TREM-2) expression tracks with M2-like macrophage activity and disease severity in COPD. Chest. 2018;153:77–86.

14. Wu K, Zhang Y, Yin-Declue H, Sun K, Mao D, Austin S, Crouch E, Brody S, Byers D, Hoffmann C, Hughes M, and Holtzman M. A correctable immune niche for basal-epithelial stem cell reprogramming and post-viral lung diseases. J Clin Invest. 2024;134:e183092.

15. Zhang Y, Mao D, Roswit WT, Jin X, Patel AC, Patel DA, Agapov E, Wang Z, Tidwell RM, Atkinson JJ, Huang G, McCarthy R, Yu J, Yun NE, Paessler SL, Lawson TG, Omattage NS, Brett TJ, and Holtzman MJ. PARP9-DTX3L ubiquitin ligase targets host histone H2BJ and viral 3C protease to enhance interferon signaling and control viral infection. Nat Immunol. 2015;16:1215–27.

16. Wu K, Zhang Y, Mao D, Iberg C, Yin-Declue H, Sun K, Keeler S, Wikfors H, Young D, Yantis J, Austin S, Byers D, Brody S, Crouch E, Romero A, and Holtzman M. MAPK13 controls structural remodeling and disease after epithelial injury. bioRxiv. 2024;10.1101/2024.05.31.596863.

17. Zhang Y, Wu K, Mao D, Iberg CA, Yin-Declue H, Sun K, Wikfors H, Keeler SP, Li M, Young D, Yantis J, Austin S, Crouch E, Chartock J, Han Z, Byers D, Brody S, Romero AG, and Holtzman M. A first-in-kind MAPK13 inhibitor that can correct stem cell reprogramming and post-injury disease. bioRxiv. 2024;10.1101/2024.08.21.608990.

18. Zhang Y, Mao D, Keeler SP, Wang X, Wu K, Gerovac BJ, Shornick LP, Agapov E, and Holtzman MJ. Respiratory enterovirus (like parainfluenza virus) can cause chronic lung disease if protection by airway epithelial STAT1 is lost. J Immunol. 2019;202:2332–47.

